# The circulating proteome and cancer risk: A systematic literature review and meta-analysis of 26 prospective studies with genetic validation

**DOI:** 10.1101/2025.04.23.25326245

**Authors:** Alison Dillman, Haige An, Zhe Huang, Wing Ching Chan, Sarah Blagden, Mahboubeh Parsaeian, Joshua R Atkins, Karl Smith-Byrne, Ruth C Travis

**Affiliations:** Cancer Epidemiology Unit, Nuffield Department of Population Health, University of Oxford, Oxford, UK; Department of Oncology, University of Oxford, Oxford, UK

**Keywords:** Systematic review, meta-analysis, proteomics, cancer, exome, Mendelian randomisation

## Abstract

**Background:** Proteins have an integral role in cancer aetiology and inform cancer detection, treatment, and prognosis. While published data from prospective proteomic studies identifying early detection cancer risk markers have rapidly increased in the past five years, the landscape of evidence remains unclear. We aim to synthesize the evidence on circulating proteins and cancer risk.

**Methods:** We conducted a systematic review and meta-analysis. We searched Embase and Medline up to December-2023 with reference-list screening and hand-searching up to June-2024. Prospective cohort studies were eligible if they used multiplex panels, included adults without cancer diagnosis at baseline, and reported associations between circulating proteins at baseline and risk of incident cancer. Based on highest global incidence, we included esophageal, stomach, colorectal, liver, lung, breast, cervical, prostate, bladder, thyroid cancer. We conducted exome-protein-score and Mendelian randomisation analyses and integrated exome-gene-burden results for protein associations that passed false-discovery-rate correction to assess protein roles in cancer aetiology.

**Findings:** Of 4,949 articles, we included 26 unique studies comprising 84,129 participants and 14,326 cases. The studies profiled 2,434 unique proteins and reported 19,130 total protein-cancer-associations. We conducted 3,448 meta-analyses and detected 216 associations (131 novel) that passed false-discovery-rate correction for stomach (n=2), colorectal (n=27), lung (n=172), breast (n=14), and prostate (n=1) cancer. No significant associations were observed for bladder cancer. Meta-analyses were not possible for esophageal, liver, cervical, or thyroid cancer, due to limited data. Supporting genetic evidence was found for 39 protein-cancer-associations.

**Interpretation:** We identified 131 novel protein-cancer-associations with strong evidence across meta-analyses of which some may be cancer markers and others may have a role in aetiology, indicated by supporting genetic analyses, including ITGA11 and lung cancer. Our findings highlight the need for large, diverse, and mature prospective proteomic cohort studies of cancer risk to ensure the equity and generalisability of insights into cancer risk.

## Panel: Research in context

### Evidence before this study

While prospective studies have mostly relied on individual or small panels of blood proteins, novel developments in multiplex proteomics methods allow the simultaneous measurement of thousands of proteins across multiple cancer endpoints. The published data from prospective proteomic studies of cancer risk has rapidly increased in the past five years. However, the current evidence is unclear regarding proteins with consistently observed associations and areas of saturation or gaps across the literature. We searched Embase and Medline from their inception to 11 December 2023 with reference-list screening and hand-searching up to 1 June 2024. Prospective cohort studies were eligible if they used multiplex panels, included adults without cancer diagnosis at baseline, and reported the association between circulating proteins at baseline and risk of incident cancer. We included esophageal (C15), stomach (C16), colorectal (C18-20), liver (C22), lung (C34), breast (C50), cervix uteri (C53), prostate (C61), bladder (C67), and thyroid (C73) cancer. We integrated genetic analyses for associations that passed false discovery rate correction, including Mendelian randomisation and exome protein score, and existing exome gene burden analyses.

### Added value of this study

To the best of our knowledge, this review is the first to synthesize the prospective proteomic evidence across common cancer sites. This study provides an overview of the current proteomic data for cancer risk, across 26 studies that included 84,129 participants and measured the association between 2,434 unique proteins and risk of ten cancers. We conducted 3,448 meta-analyses and report 216 proteins associated with risk of at least one of the ten cancers, of which 131 were novel associations, with genetic results providing support for the role of several proteins in cancer aetiology.

### Implications of all the available evidence

Our findings have several potential implications for future research and clinical practice. We find prospective evidence together with the new genetic data that implicate circulating proteins in relation to cancer aetiology, such as ITGA11 and lung cancer. We also find consistent evidence across multiple independent studies for previously reported candidates for early cancer detection, such as GDF15 and risk of stomach and colorectal cancer, as well as multiple strong protein associations with lung cancer risk including CEACAM5. However, most available evidence was for proteins and cancer risk in white participants and there was insufficient evidence for meta-analyses of multiple common cancer sites including the liver and esophagus. Therefore, to deliver on the potential promise of plasma proteomics for cancer research, there is a need for large and mature prospective proteomic studies in diverse cohorts with high throughput methods. The capacity to look across the blood proteome more widely provides greater insights into pathways that may be important in carcinogenesis and may ultimately inform studies of potential therapeutic interventions for cancer prevention.

## Introduction

Cancer is a leading cause of morbidity and death, with an estimated 20 million new cases and 9·7 million deaths globally in 2022.^1^ A 77% increase in the burden of cancer is predicted by the World Health Organization by 2050, to reach over 35 million new cases annually.^1^ Although a limited set of assays are currently used in standard clinical practice to measure circulating proteins like PSA, CA125, CEA, less than 20% of all cancers have reliable biomarkers to assist in their early detection. There is a critical need to better understand cancer aetiology to identify new protein-based biomarkers that can assist in the early detection of cancer and provide opportunities for prevention and early detection.

Levels of circulating proteins reflect human biological processes including cell growth, proliferation, dysregulation, and apoptosis. Prior studies have identified the role of proteins in cancer aetiology. For instance, IGF-I can be measured in the circulation and elevated levels have been associated with increased risk of breast, colorectal and prostate cancer.^2^ Similarly, circulating levels of MSMB are inversely associated with risk of prostate cancer.^3^ However, associations between most circulating proteins and cancer risk remain unknown.

While most prospective studies have relied on individual or small panels of blood proteins, novel developments in multiplex proteomics methods allow the simultaneous measurement of thousands of proteins across multiple cancer sites.^4^ The published data from prospective proteomic studies of cancer risk has rapidly increased in the past five years. However, the literature remains unclear and there is a need to synthesize the evidence, identify gaps in the research, and inform future directions.^5^

## Methods

### Search strategy and selection criteria

The Preferred Reporting Items for Systematic Reviews and Meta-Analyses (PRISMA) guidelines were followed,^6^ which require transparent reporting of the study purpose, methods, and findings. The preliminary purpose and methods were presented 21-November-2023 at the European Prospective Investigation into Cancer and Nutrition (EPIC)-Oxford Participant Panel for input and engagement by participants in a current prospective proteomic cohort study.^7^ Embase and Medline were searched up to 11-December-2023. Reference lists were reviewed for articles included in the final selection and hand-searches were conducted up to 1-June-2024. Search terms are reported in *Supplementary Table 1*.

Full-text peer-reviewed publications of prospective cohorts with baseline blood biomarker assessment and multiplex protein panels (i.e., panels measuring >1 protein) were eligible for inclusion. Studies reporting associations between circulating protein levels measured at baseline and incident cancer diagnosis after follow-up as relative risk, odds ratio, or hazard ratio were accepted for inclusion. Proteins identified as associated with subsequent cancer diagnoses are referred to here as protein-cancer-associations. Participants were required to be aged 18 years and older without a cancer diagnosis at baseline. Based on highest global incidence, cancers of the esophagus(C15), stomach(C16), colorectum(C18-20), liver(C22), lung(C34), breast(C50), cervix uteri(C53), prostate(C61), bladder(C67), and thyroid(C73) were included.^1^ No date or language restrictions were applied.

Two reviewers (AD, HA) independently screened titles and abstracts of retrieved articles. These same two reviewers obtained full-text publications for relevant abstracts and independently screened each publication against the eligibility criteria. Disagreements were discussed with a third reviewer (KSB, RT) to reach consensus. This review is registered at PROSPERO, CRD42023482553.

### Statistical analysis

Study characteristics, participant characteristics, and multivariable-adjusted effect estimates for each study were extracted by the reviewers. When multiple publications reported on the same cohort and cancer site, the data for the protein-cancer-association with the largest sample size and longest follow-up was extracted as the primary source. Authors were contacted to request missing data. Methodological quality was assessed using the Newcastle-Ottawa Scale for observational studies.^8^

Inverse variance-weighted fixed-effect meta-analyses were conducted for each protein-cancer-association reported in two or more studies.^9^ Subgroup and sensitivity analyses were not conducted due to the low number of studies per protein-cancer-association. The log of hazard ratio (HR), odds ratio (OR), or relative risk (RR) per change in standard deviation (SD) in protein level with 95% confidence interval (CI) was used to compute combined effect estimates of protein levels and cancer risk.

The Cochran Q statistic^10^ and *I*^2^ statistic^11^ were used to calculate the percentage of observed variability due to study heterogeneity. Due to the low number of studies for each cancer site and protein, it was not feasible to generate funnel plots and perform Egger’s linear regression tests to investigate reporting biases for all outcomes.^12^ Analyses were conducted using R version 4.3.2. False-discovery-rate (FDR) adjustment was applied to all associations to account for multiple comparisons with an adjusted p<0·05 deemed statistically significant.

To further investigate and contextualize results for FDR-significant circulating protein-cancer-associations, we conducted cis-pQTL Mendelian randomisation (MR) with colocalization and exome protein scores using the UK Biobank.^13^ Further details are available in the *Supplementary Methods*. Additionally, we integrated recent exome gene burden results from a recent international meta-analysis.^14^ MR provides evidence regarding causal inference between modifiable variables and disease through using the existence of genetic variants as natural experiments, and is thus less likely to be affected by confounding and reverse causality.^15^ Colocalization assesses whether there is a shared genetic signal between multiple variants.^16^ The genetic methods provide insight into consequences of higher protein levels throughout life, with loss-of-function providing insight into disruptions to protein-coding genes and lower protein levels throughout life.

## Results

Of 4,849 articles identified in the search strategy, 502 duplicates were excluded, 4,149 articles were excluded after title and abstract screening, and 168 articles were excluded after full-text screening. Overall, 30 articles representing 26 unique studies met the inclusion criteria (*Figure 1)*.^4,17–46^ Risk of bias of included studies was considered moderately low overall (*Supplementary Table 2*).

**Figure 1.**
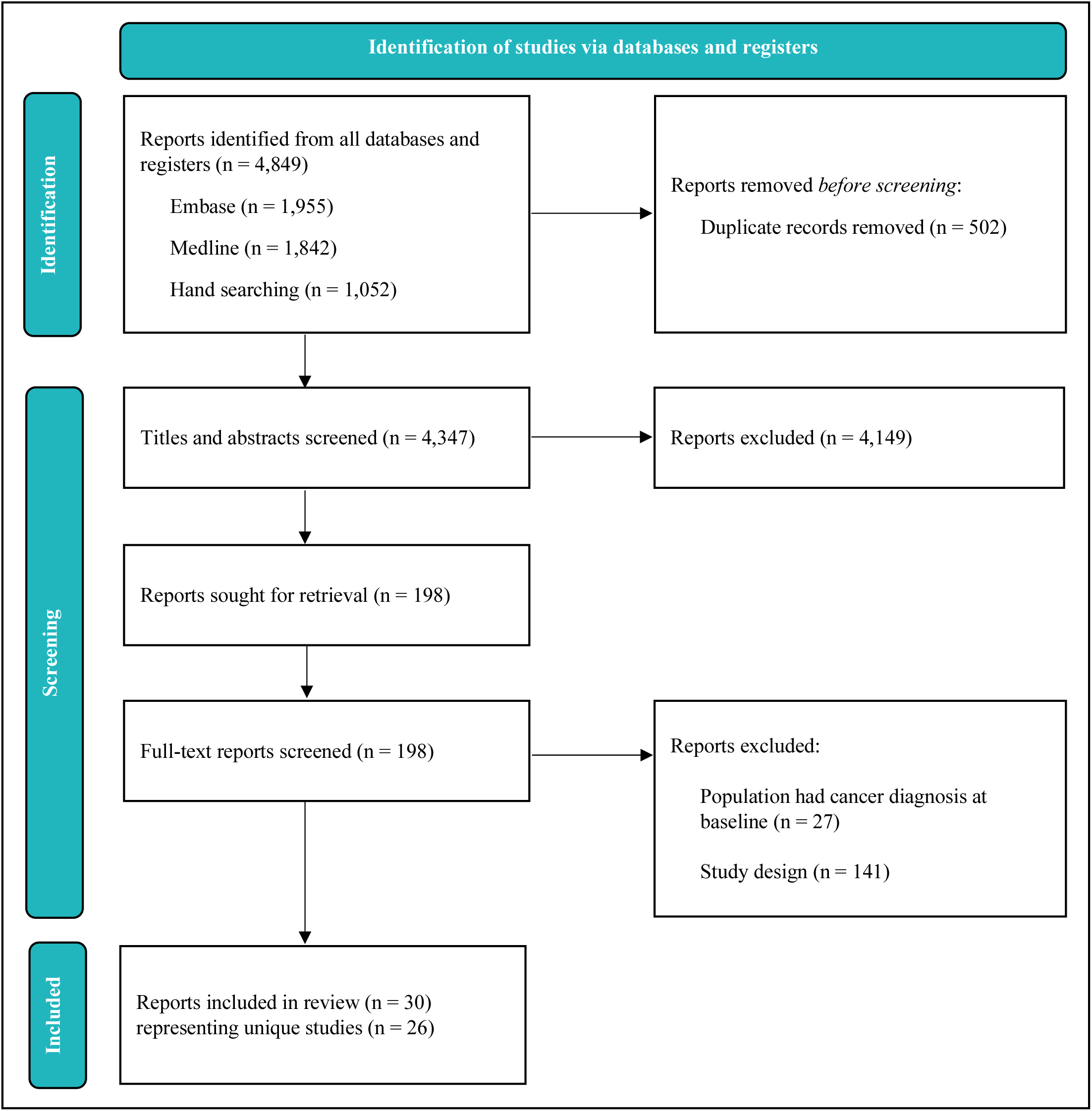
Study selection flowchart.

*Table 1* and *Figure 2* show the main characteristics of included studies. Eighteen publications were based on results from nested case-control studies, four were case-cohort studies and four were cohort studies. All studies used baseline questionnaires and blood samples. Each study profiled up to 2,204 proteins using multiplex technologies including Luminex (15 studies), Olink (n=7), mass spectrometry (n=2), Somalogic (n=1), and Linco-3-plex (n=1). Sample sizes ranged from 96 to 44,645 participants (median:758, interquartile range:869). The studies were conducted across North America (n=11), Asia (n=10), and Europe (n=8). Studies were predominantly conducted in white (85·3%) participants, with fewer studies including Asian (6·7%), black (1·4%), Hispanic (0·4%), or other (6·2%) participants. Across all 84,129 participants, 54·9% were female and 45·1% were male, with median age at baseline ranging from 47·1 to 66·4 years.

**Figure 2.**
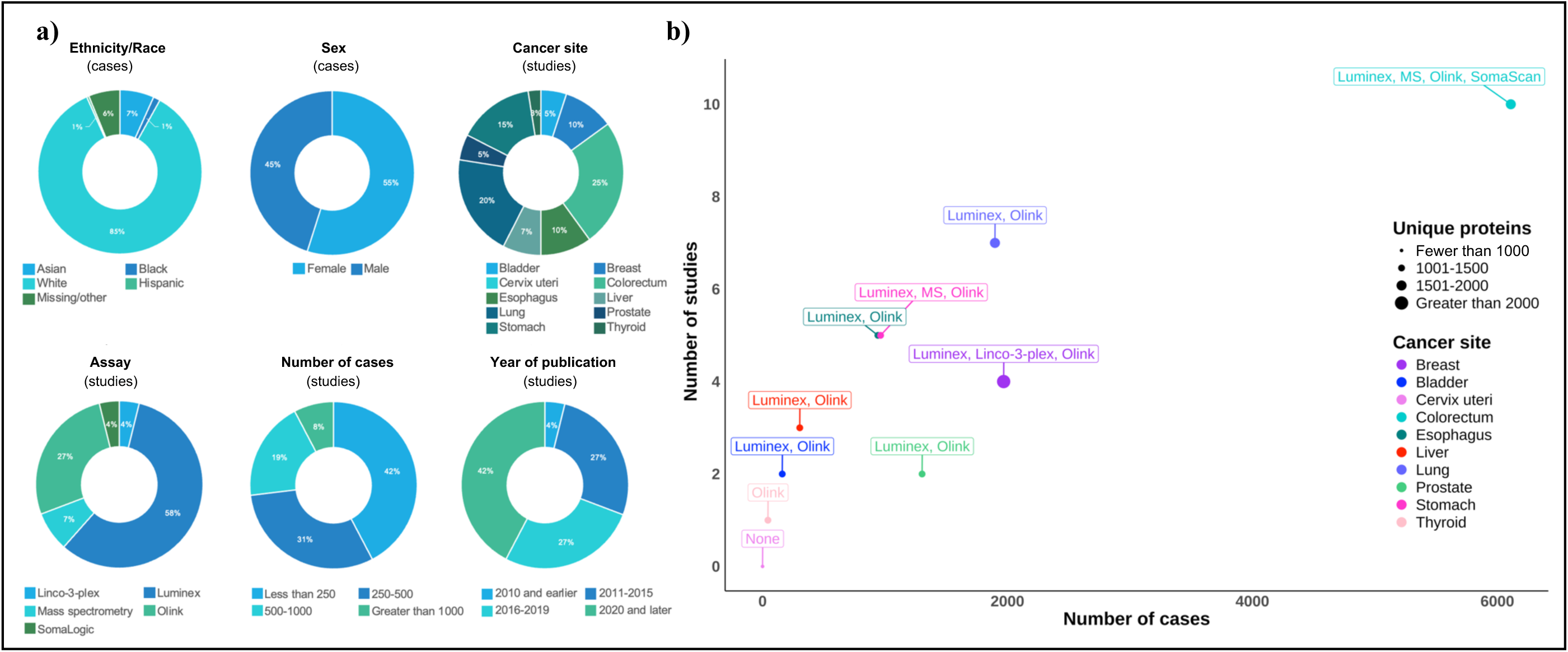
Study and participant characteristics of included cohorts. **Abbreviations**: MS, mass spectrometry. **Notes:** Percentages reflect the number of cases or studies as stated.

**Table 1.**
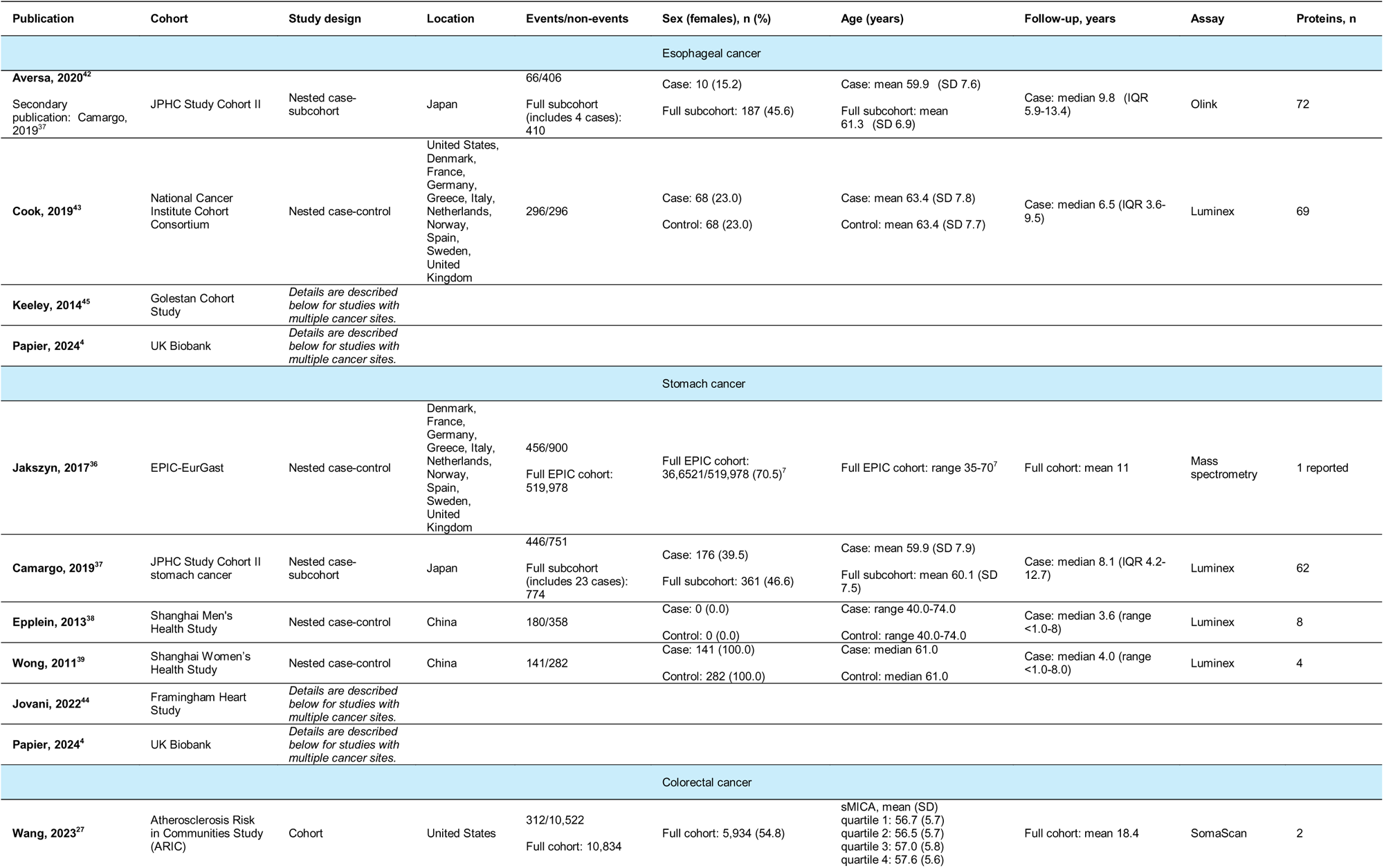
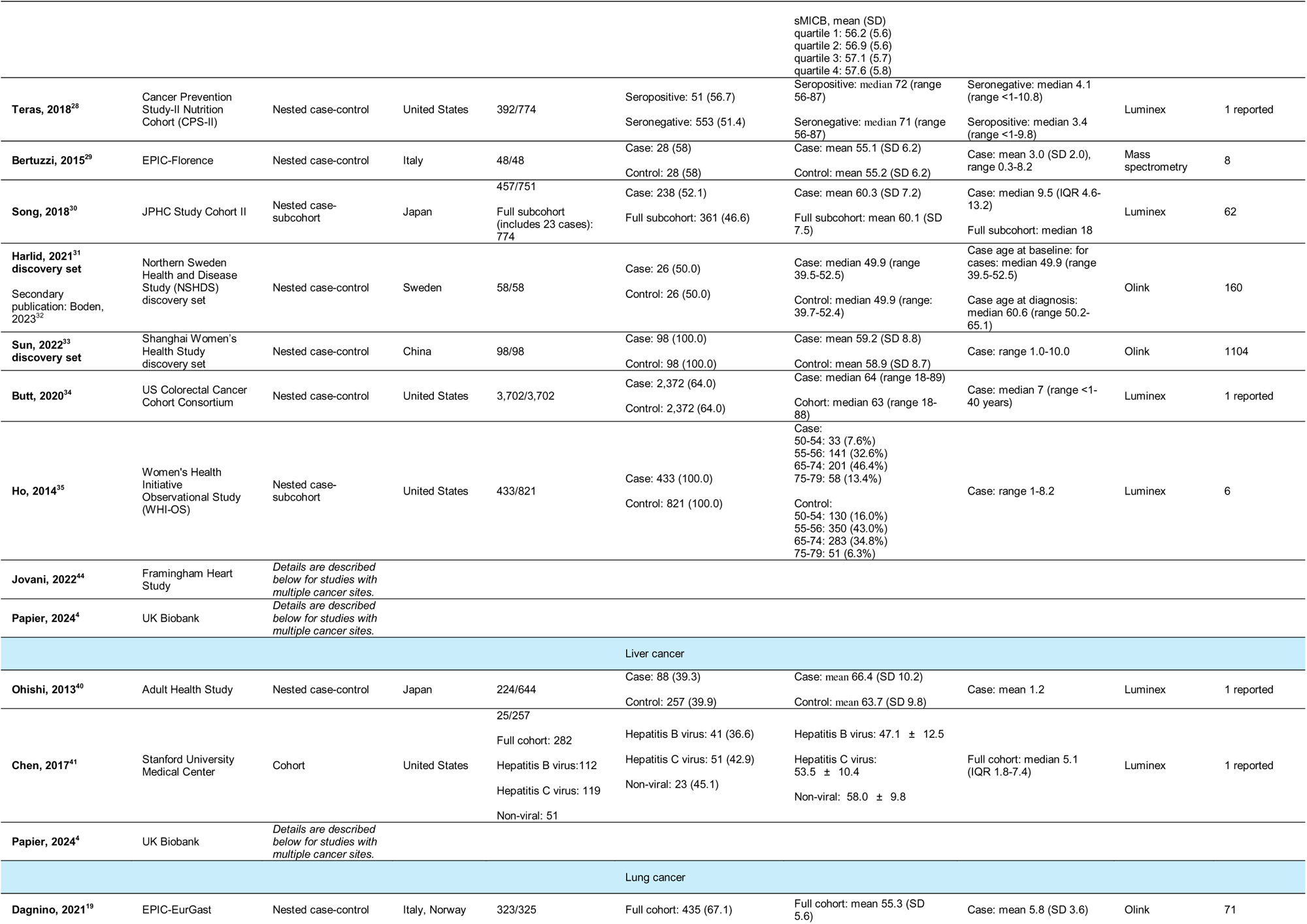
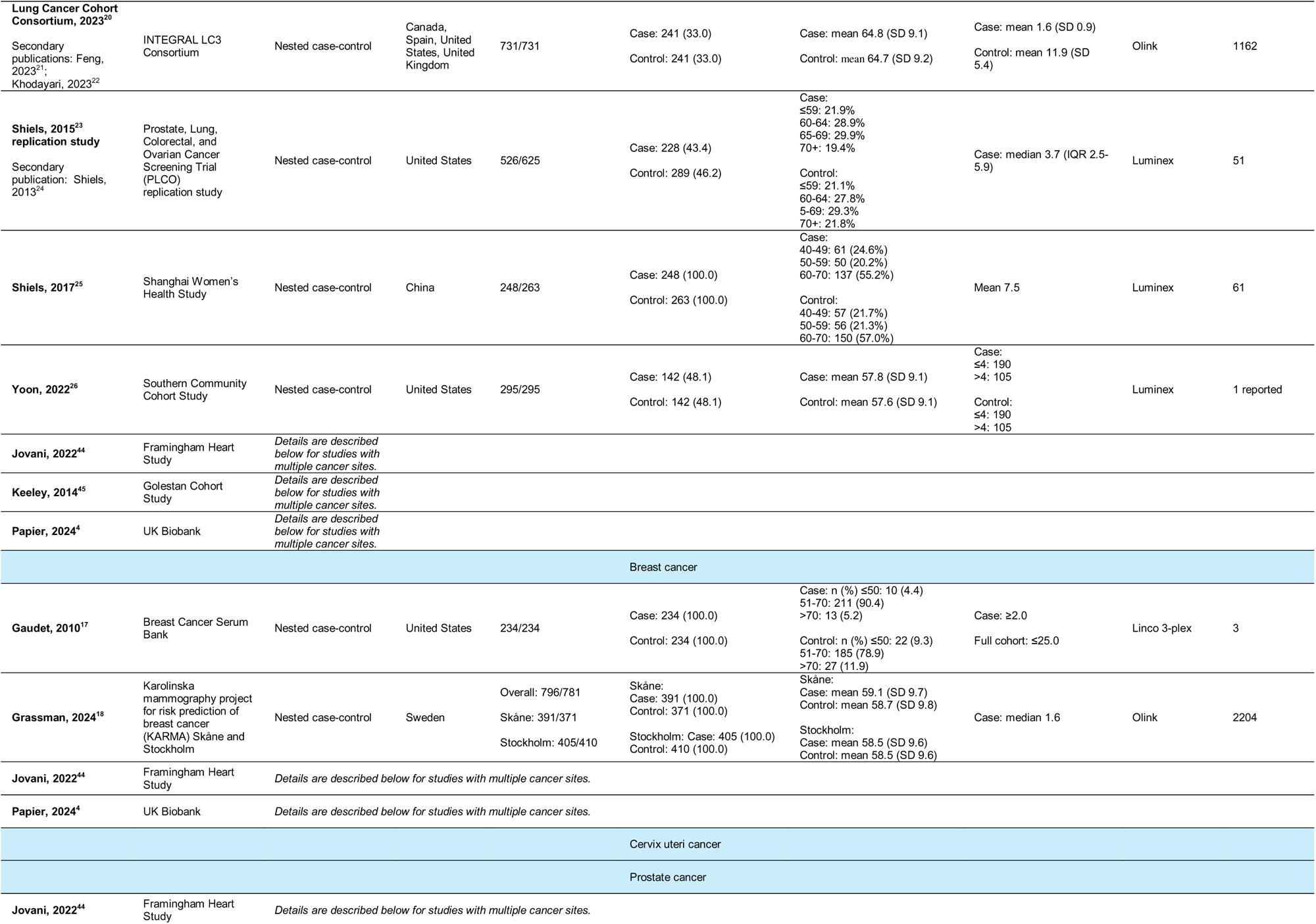
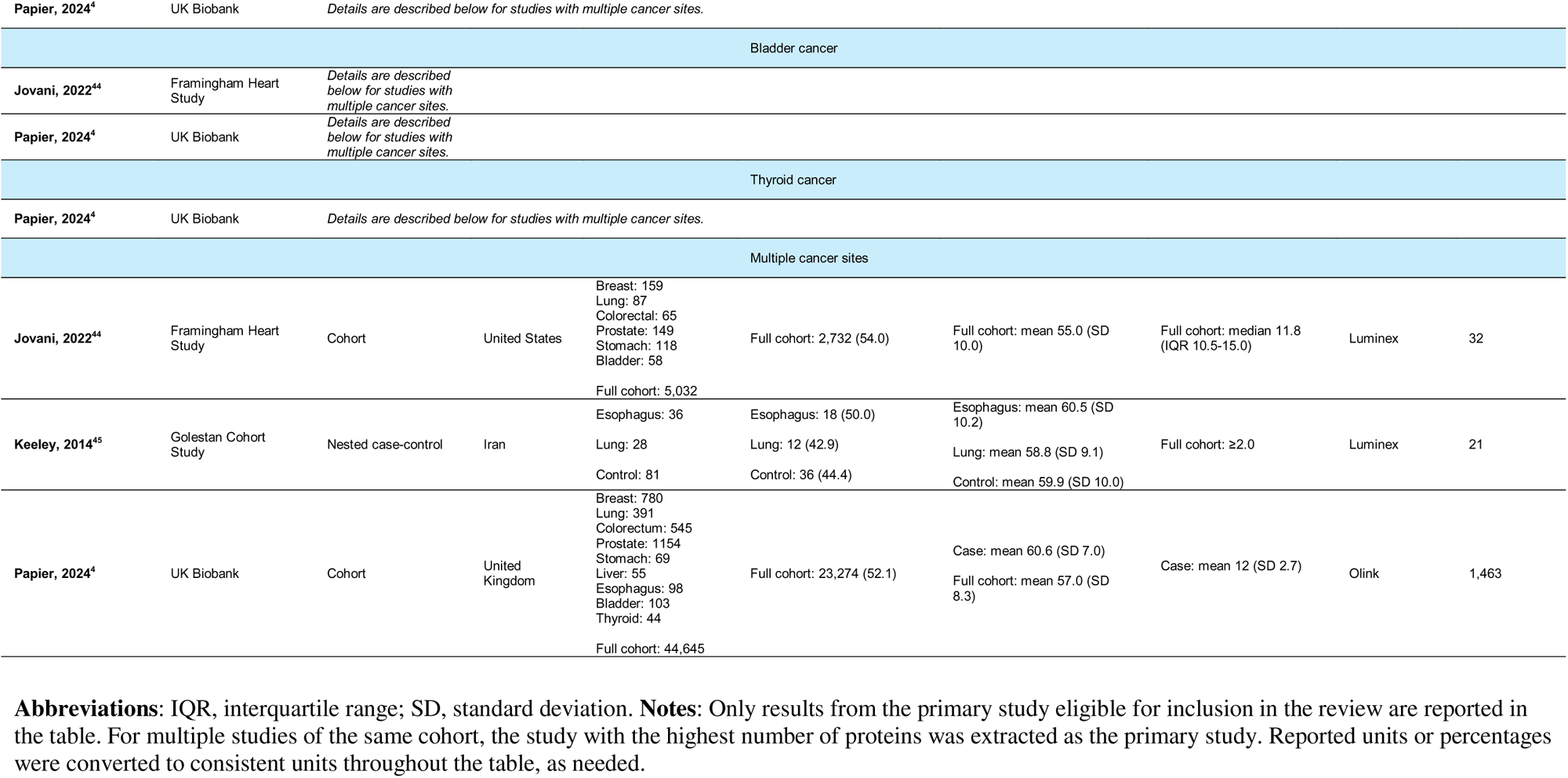
Main characteristics of included studies.

Over mean follow-up times from 1·6 to 12 years until diagnosis, 14,326 participants who developed cancer were compared to 69,803 participants without cancer diagnoses. All studies used clinical diagnoses or International-Classification-of-Disease (ICD) to identify cases, while five also allowed for self-report verified with medical record review. The number of cases included in studies ranged from 25 for liver to 3,702 for colorectal.

A total 19,137 protein-cancer-associations were investigated across nine sites for 2,434 unique proteins across all studies, including esophagus (4-studies; 1,477 unique proteins), stomach (6-studies; 1,483 unique proteins), colorectum (10-studies; 1,531 unique proteins), liver (3-studies; 1,464 unique proteins), lung (8-studies; 1,531 unique proteins), breast (4-studies; 2,366 unique proteins), prostate (2-studies; 1,463 unique proteins), bladder (2-studies; 1,464 unique proteins), and thyroid (1-study; 1,463 unique proteins). No studies investigated cervix uteri.

Protein-cancer-associations were investigated in protein measurement-specific quantiles or other categories for 143 unique proteins across six sites, including the esophagus, stomach, colorectal, liver, lung, and breast. The associations included sICAM-1 (HR:4·42, 95%CI:1·84-10·61; above vs. below mean) with hepatocellular carcinoma^41^, and sIL-1R2 (HR:0·44, 95%CI:0·29-0·67; quartile 4 ≥9·2ng/mL vs. quartile 1 <5·8ng/mL) with colorectal.^35^

Protein-cancer-associations were reported in single studies only for 10,776 associations across 2,446 unique proteins (*Supplementary* Figure 1). Observed associations included TSPYL1 (HR:0·79, 95%CI:0·69-0·90)^18^, VTCN1 (HR:1·12, 95%CI:1·05-1·19)^4^, STX6 (HR:0·87, 95%CI:0·80-0·95)^4^ with breast, WAS (HR:0·54, 95%CI:0·40-0·73)^4^, ICAM4 (HR:0·75, 95%CI:0·61-0·91), S100A16 (HR:1·28, 95%CI:1·11-1·47) with bladder^4^, and TFF1 (HR:1·19, 95%CI:1·10-1·30), RBP2 (HR:1·19, 95%CI:1·09-1·30), SPINK4 (HR:1·19, 95%CI:1·09-1·30) with colorectal.^4^ REG4 (HR:1·83, 95%CI:1·53-2·20), CCL14 (HR:1·33, 95%CI:1·17-1·51), ST6GAL1 (HR:1·36, 95%CI:1·18-1·55) were associated with esophageal,^4^ and IGFBP3 (HR:0·46, 95%CI:0·39-0·54), IGFBP7 (HR:1·65, 95%CI:1·48-1·84), BST2 (HR:1·72, 95%CI:1·52-1·94) with liver.^4^ IFI30 (OR: 1·49, 95%CI:1·30-1·70)^20^, TREM2 (HR:1·32, 95%CI:1·19-1·47)^4^, SFTPA2 (HR:1·24, 95%CI:1·14-1·35)^4^ were associated with lung. GP2 (HR:1·29, 95%CI:1·21-1·36), TSPAN1 (HR:1·14, 95%CI:1·10-1·18), FLT3LG (HR:0·87, 95%CI:0·82-0·92) were associated with prostate cancer risk.^4^ ANXA10 (HR:1·75, 95%CI:1·51-2·02), TFF1 (HR:1·90, 95%CI:1·58-2·28), GGH (HR:1·58, 95%CI:1·35-1·86) were associated with stomach^4^ and APBB1IP (HR:0·46, 95%CI:0·30-0·69), CAPG (HR:0·57, 95%CI:0·42-0·78), KLK8 (HR:1·54, 95%CI:1·21-196) were associated with thyroid.^4^ Of note, ENPP2 (HR:2·65, 95%CI:2·11-3·34), NFASC (HR:2·49, 95%CI:1·97-3·14), and CDHR2 (HR:2·77, 95%CI:1·94-3·97) protein-liver cancer associations were among the highest reported relative risks across all cancer sites (*Supplementary* Figure 1).^4^ Only one study reported each unique protein-cancer-association for esophageal, liver, and thyroid. Thus, meta-analyses were not possible for the respective cancer sites given that meta-analysis required a minimum of two studies for the same protein-cancer-association.

A total of 3,448 meta-analyses were conducted across all studies, including stomach (2-studies; 7 unique proteins), colorectum (4-studies; 1,022 unique proteins), lung (4-studies; 1,114 unique proteins), breast (3-studies; 1,301 unique proteins), prostate (2-studies; 2 unique proteins), and bladder (2-studies; 2 unique proteins). Of the 3,448 protein-cancer-associations, 560 were nominally significantly associated with cancer risk (*Figure 3, Figure 4*). After applying FDR-correction, 216 protein-cancer-associations were significant, of which 176 were associated with an increased risk, and 40 were associated with a lower risk. Notably, 131 of the associations identified were not previously significantly associated in comprising studies for the same endpoint. FDR-significant heterogeneity (Q_p-FDR_≤0·05) was observed among studies for seven of the FDR-significant protein-cancer-associations, for which observed heterogeneity between the studies ranged from *I*^2^=90·26% to *I*^2^=96·49%. Given the low number of included studies, *I*^2^ statistic and Q have lower precision and power, consequently these should be interpreted with caution.^47^ Across all meta-analyses, the number of cases ranged from 156 to 1,390.

**Figure 3.**
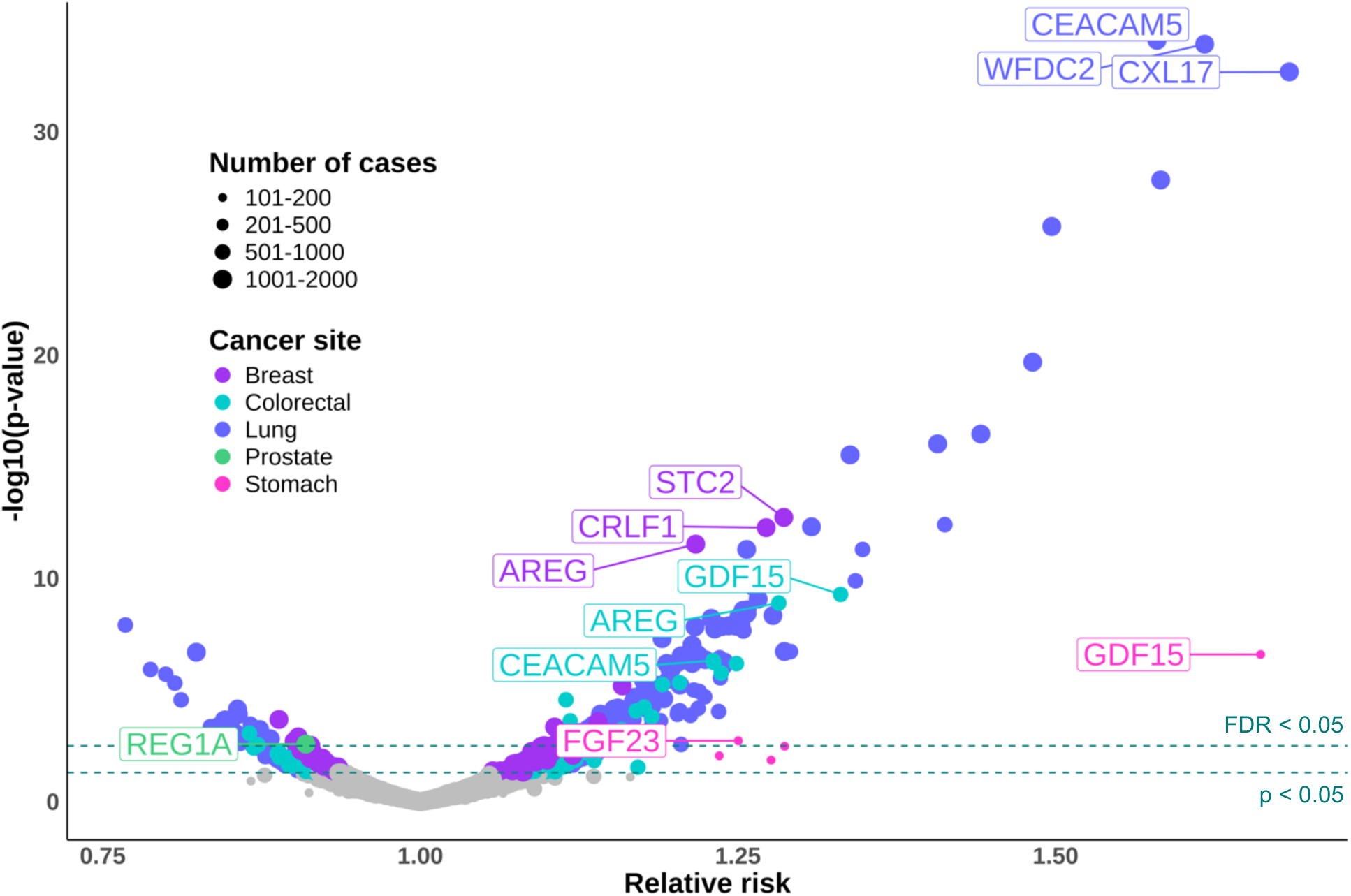
Associations of circulating proteins and cancer risk from the 3,448 meta-analyses by cancer site. **Abbreviations**: FDR, false discovery rate. **Notes**: Each point represents a meta-analysis. The size of each point corresponds to the number of cases and the color of each point corresponds to the cancer site. The top three FDR-significant protein-cancer-associations are labelled for each cancer site.

**Figure 4.**
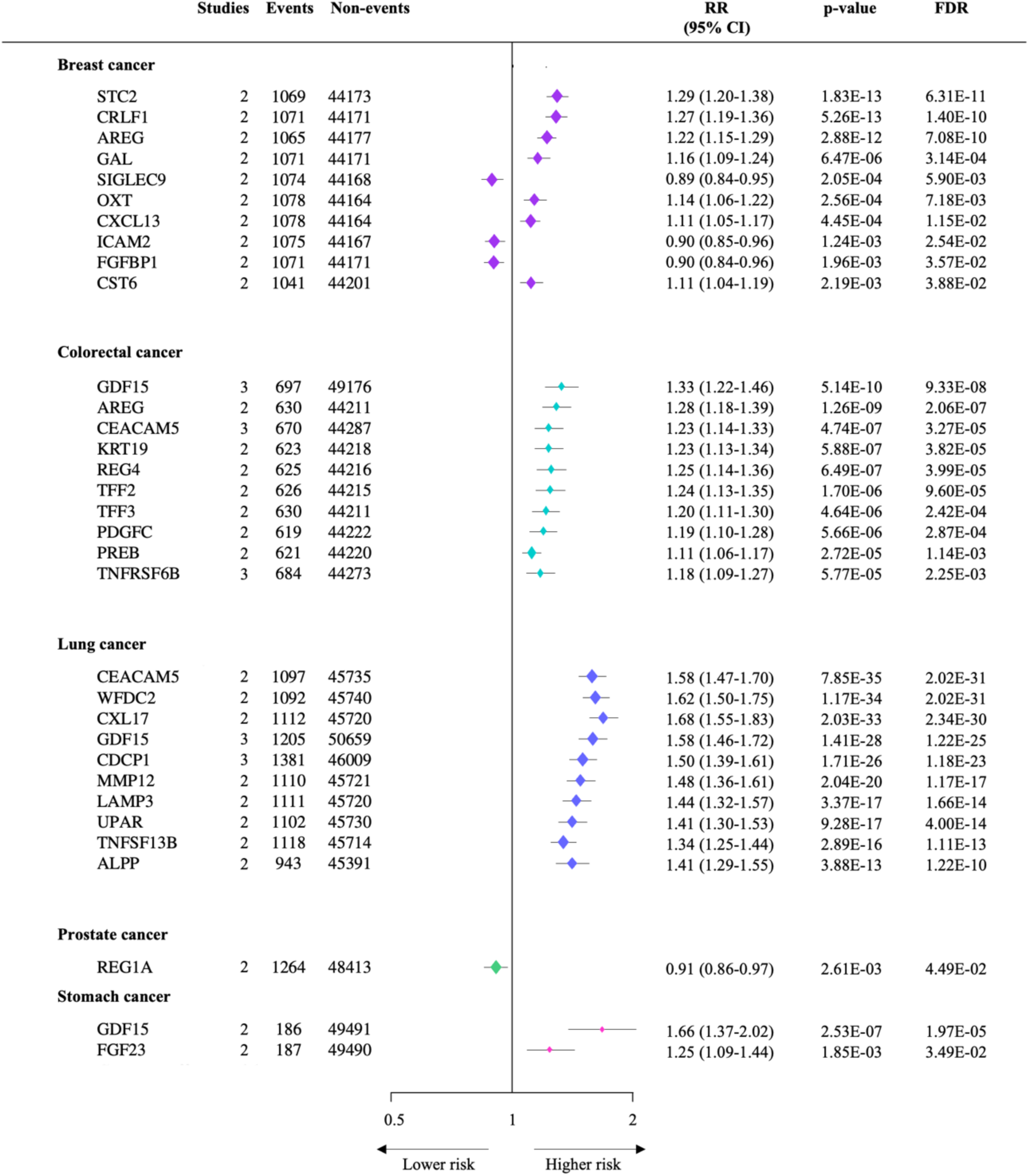
Relative risks (95% confidence intervals) for selected associations between circulating proteins and cancer risk from meta-analyses of study-specific results from up to 3 prospective studies. **Abbreviations**: CI, confidence interval; FDR, false discovery rate; RR, relative risk. **Notes**: All protein-cancer-associations are FDR-significant (<0.05) and presented in ascending order (strength of association) by cancer site. Results are shown for up to ten FDR-significant associations for each cancer site from the meta-analyses. RRs were combined using inverse-variance-weighted averages of the log RRs in the separate studies, yielding a result and its 95%CI, which is plotted as a diamond, with area inversely proportional to the variance of the log RR.

Across 1,301 meta-analyses conducted for breast cancer risk (mean 2·0 studies; 522-1,231 cases), 14 proteins remained significant after FDR-correction, including those with strong positive associations, STC2 (relative risk per SD increment (RR): 1·29, 95%CI:1·20-1·38), CRLF1 (RR:1·27, 95%CI:1·19-1·36), AREG (RR:1·22, 95%CI:1·15-1·29). In contrast, SIGLEC9 (RR:0·89, 95%CI:0·84-0·95), ICAM2 (RR:0·90, 95%CI:0·85-0·96), FGFBP1 (RR:0·90, 95%CI:0·84-0·96), TYRO3 (RR:0·91, 95%CI:0·85-0·97), TSHB (RR:0·91, 95%CI:0·86-0·97) were associated with a lower breast cancer risk.

Of 1,114 meta-analyses conducted for lung cancer (mean 2·1 studies; 399-1,390 cases), 172 FDR-significant protein-cancer-associations were detected. Among these, CEACAM5 (RR:1·58, 95%CI:1·47-1·70), WFDC2 (RR:1·62, 95%CI:1·50-1·75), CXCL17 (RR:1·68, 95%CI:1·55-1·83) had strong positive associations with lung cancer risk. Conversely, 32 of the 172 proteins were associated with a lower lung cancer risk, including TNR (RR:0·77, 95%CI:0·70-0·84), SCF (RR:0·82, 95%CI:0·77-0·89), and GDF8 (RR:0·79, 95%CI:0·72-0·87).

Across 1,022 meta-analyses conducted for colorectal cancer (mean 2·1 studies; 156-757 cases), 27 FDR-significant protein-cancer-associations were detected. Of these, GDF15 (RR:1·33, 95%CI:1·22-1·46), AREG (RR:1·28, 95%CI:1·18-1·39) and CEACAM5 (RR:1·23, 95%CI:1·14-1·33) had strong evidence for a positive association with colorectal cancer risk. In contrast, associations with a lower colorectal cancer risk were observed for DPP4 (RR:0·87, 95%CI:0·80-0·94), ITGA11 (RR:0·87, 95%CI:0·80-0·95).

Seven meta-analyses for stomach cancer were conducted (2 studies; 185-187 cases), of which associations with two proteins remained significant after FDR-correction, GDF15 (RR:1·66, 95%CI:1·37-2·02) and FGF23 (RR:1·25, 95%CI:1·09-1·44). Two meta-analyses were possible for prostate (2 studies; 1,264-1,266 cases), of which REG1A (RR:0·91, 95%CI:0·86-0·97) was associated with a lower prostate cancer risk after FDR-correction. Of the two meta-analyses conducted for bladder (2 studies; 159-161 cases), no significant protein-cancer-associations were observed.

Several of these protein-cancer-associations were observed (both FDR-significant and directionally concordant) across more than one site. For example, AREG was positively associated with breast, lung, and colorectal cancer risk. Similarly, TFF3 was positively associated with both colorectal and lung cancer risk. GDF15 and FGF13 were positively associated with stomach, lung, and colorectal cancer risk, and CXCL13 was positively associated with both lung and breast cancer risk.

We sought to integrate additional genetic evidence for observed protein-cancer-associations to triangulate the potential aetiological role of proteins in cancer risk. To do so, novel genetic analyses were integrated for the 216 proteins (156 using cis protein quantitative trait loci [pQTL] exome analyses, 173 using cis pQTL MR analyses) across all sites that survived FDR-correction in the meta-analyses of prospective data. Of these, seven conventionally significant protein-cancer-associations (4 concordant, 3 discordant) were found in exome analyses and 33 were found in MR analyses (12 concordant, 21 discordant) (*Figure 5, Supplementary* Figure 2). Directionally concordant exome evidence was found for four proteins (CEACAM5, ALPP, VEGFA, EDA2R) associated with increased lung cancer risk. Directionally concordant MR evidence was found for 12 proteins, including five each for colorectal (TFF3, ADM, TNFRSF6B, PI3, MMP12) and lung (TIE1, ICAM4, SFRP1, ITGA11, COMP), and two for breast (CST6, APOH). A discordant direction of effect was found for three proteins (SPINK1-colorectal, CEACAM5-colorectal, CLEC4D-lung) in exome analyses, and 21 proteins in MR analyses (including COL4A1-lung, FCAR-lung, FGF21-colorectal). CEACAM5 was associated with a lower colorectal cancer risk across both exome and MR analyses, while observational evidence showed an association with a higher risk. Of the 12 concordant proteins, colocalization data for signals <5×10^−05^ were only available for TNFRSF6B, for which an association was observed with prostate cancer.^16^ However, colocalization is not particularly stable if there are not strong signals in both endpoints and small sample size further limits statistical power.^16^

**Figure 5.**
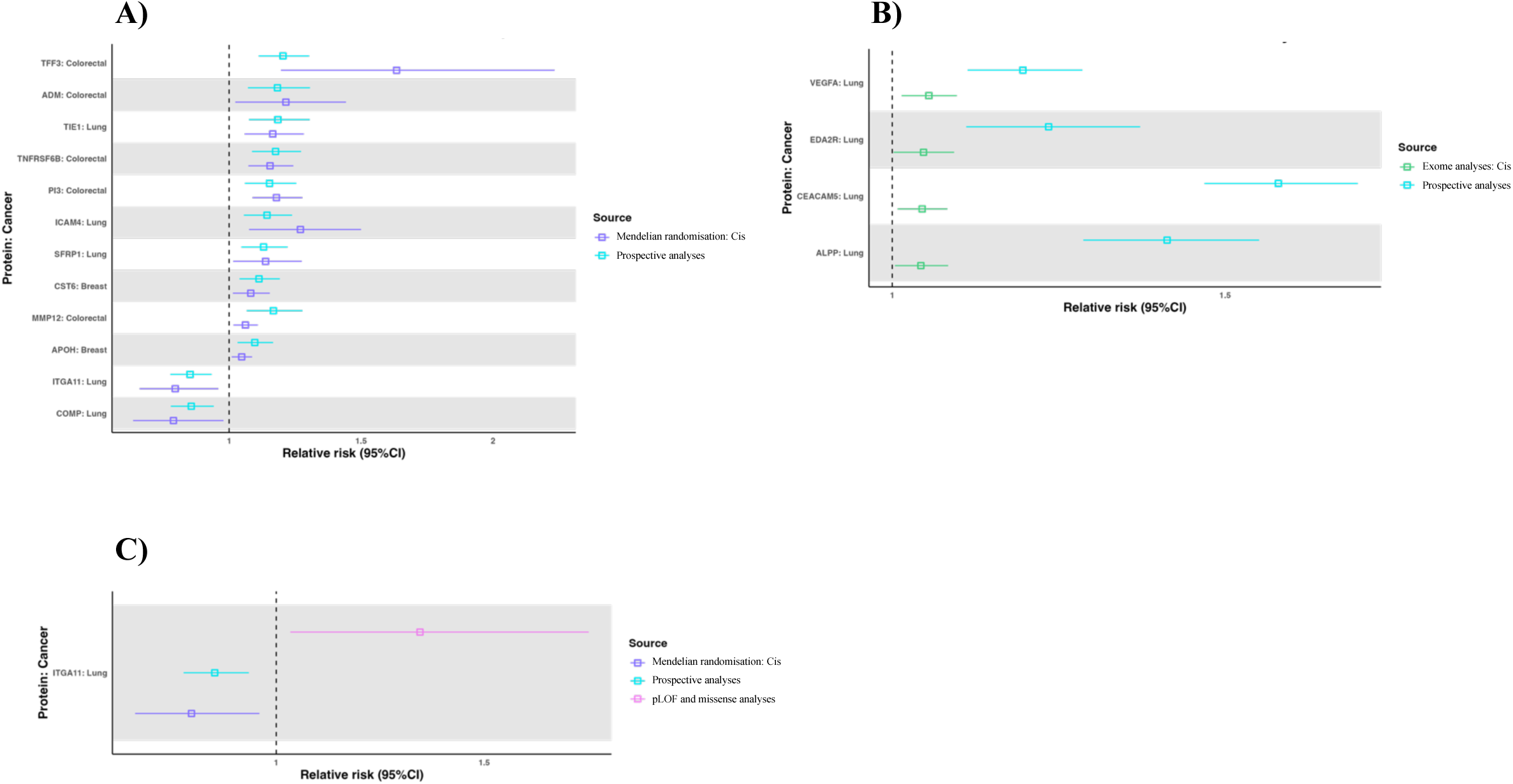
Genetic evidence for proteins identified in the meta-analyses of prospective data: Concordant evidence compared to the observational analyses. Mendelian randomisation (A) and exome (B) evidence concordant with the observational analyses; C) Probable loss-of-function (pLOF) and missense analyses evidence for protein-cancer-associations. **Abbreviations**: CI, confidence interval; FDR, false discovery rate; pLOF, probable loss-of-function. **Notes:** Relative risks (95% confidence intervals) for proteins identified in the meta-analyses of prospective data, for which genetic evidence was also available and directionally concordant associations were observed.

Additionally, results were extracted from a recent large meta-analysis of probable loss-of-function (pLOF) and missense analyses.^14^ Thirty-six of the 39 proteins with cis MR and exome evidence were reported in the pLOF and missense analyses (EDA2R-lung, SPON1-lung, MMP12-colorectal were not available). Of these, eight were conventionally significant (*Figure 5, Supplementary* Figure 2). Five (ICAM1, MAD1L1, COL4A1, ST6GAL1, ITGA11) associated with lung and one (TFF3) associated with colorectal cancer risk were conventionally significant in pLOF and missense analyses. One (CREG1-breast) was conventionally significant in missense analyses and one (ALPP-lung) was conventionally significant in pLOF analyses. Of the three proteins not reported in the pLOF and missense analyses, one (EDA2R-lung) was found in Genebass however the results were not significant. Concordant evidence was observed for the association between ITGA11 and lung cancer risk, wherein both higher observed and genetically predicted levels of the protein were associated with a reduced risk, and loss-of-function was also associated with an increased risk (RR:1·35, 95%CI:1·03-1·75).

## Discussion

This study provides an overview of the current proteomic evidence for cancer risk, across 26 studies that included 84,129 participants and measured the association between 2,434 unique proteins and risk of ten common cancers. Across the 3,448 meta-analyses, 216 proteins were associated with risk of at least one of the ten cancers, of which 131 novel protein-cancer-associations were identified that had not previously been reported.

Significant protein-cancer-associations were observed across cancer sites, including lung and colorectal for circulating levels of CEACAM5, a cell-surface glycoprotein that promotes cell proliferation and migration, implicated in tumorigenesis and metastasis.^48,49^ Similarly, circulating GDF15, a protein from the transforming growth factor β superfamily and implicated in the development and progression of several cancers,^50^ was associated with lung, colorectal, and stomach. STC2 was strongly associated with breast, which is consistent with previous evidence implicating the protein in breast cancer aetiology, through pathways of cell proliferation and metastasis, and as a possible therapeutic target.^51^ The growth factor REG1A was a novel finding associated with a lower prostate cancer risk in the meta-analysis, while SIGLEC9, ICAM2, and FGFBP1 were associated with a lower breast cancer risk, which may suggest a protective role of the proteins in cancer aetiology.

While associations that we identify may be explained as potentially modifiable risk factors through mechanisms such as inflammation or growth factors, proteins may also be secreted by tumours, or represent the body’s response to a yet undiagnosed cancer and provide evidence for potential early detection proteins.^52^ Strong and consistent evidence was found for markers that may be less likely to be aetiological but may have utility in the context of early cancer detection, for example known biomarkers CEACAM5,^49^ WFDC2,^53^ and CXCL17^20^ with lung cancer. The clinical impact of such findings extends across opportunities for precision prevention, surrogate markers, early detection, and potential therapeutic targets.

Notably, ITGA11 was protective for lung cancer across prospective and genetic analyses, with loss-of-function (i.e., the inheritance of suppressed or non-functioning copies of the protein) associated with an increased risk, supporting the rationale of a protective role within cancer aetiology. ITGA11 was not significant across comprising observational studies, however the association was FDR-significant once meta-analysed across 632 cases, highlighting the value of pooling data for greater statistical power. While prior research has found the overexpression of ITGA11 in lung cancer and an association with lower recurrence-free survival,^54^ this may be explained by increased cellular expression in the presence of a subclinical tumour. The novel genetic analyses we report here provide new insight into ITGA11 as a possibly protective protein, as increased levels of ITGA11 may be elevated in response to a tumour. Conversely, TFF3, ADM, VEGFA, and EDA2R, among others, were associated with an increased colorectal or lung cancer risk across the observational and genetic analyses, providing evidence to support the proteins as possible risk factors. Of note, where protein-cancer-associations were not observed in the genetic data may also be explained by a lack of strong instrument or underpowered analyses.^5^

This study reveals a concerning lack of diversity across global proteomic data, with published prospective data being from predominantly white European participants. Such a lack of representation means it was not possible to explore the impact of ethnicity on protein-cancer-associations and the generalizability of results to underrepresented groups cannot be assumed.^55^ Few studies have investigated the potential heterogeneity of protein-cancer-associations by ethnicity but there is evidence that the inherited determinants may differ importantly between ancestries.^4,56–59^ An inequitable landscape of evidence generation weakens opportunities to fully understand and target protein-cancer-associations across diverse populations and reinforces global disparities in health. The study also highlights a paucity of data for cancer sites other than the most common, with few relatively few data available for esophagus, liver, and thyroid, for which meta-analyses were not possible, and cervix uteri, for which no eligible studies were found. Notably, the global incidence of cervical cancer is highest among countries with low socioeconomic status and poor access to early screening and prevention,^60^ across populations that have not been included in current proteomic studies. Among the top ten highest-incident cancer sites,^1^ these striking gaps in the literature represent important and impactful areas for future research.

To the best of our knowledge, this review is the first to synthesize the prospective proteomic evidence across common cancer sites. Strengths include a comprehensive search strategy and thus a large amount of pooled data that allowed us to assess protein-cancer-associations with greater statistical power than in single studies. A robust approach was taken with complementary genetic analyses to contextualize the results and further investigate cancer aetiology. These data provide insight into the association between proteins and cancer risk based on the current evidence, in addition to research gaps and areas of saturation across the proteomic field. There is a need for extensive prospective validation of promising candidates with conversion to easily scalable methods, for example ELISA, that can be reliably used in clinical settings.

The study has limitations. The review is limited to published evidence. Consequently, meta-analyses with fewer participants had lower statistical power and subgroup and sensitivity analyses were not possible. The review focused on studies that used proteomic panels to assess the landscape of evidence in the era of rapidly evolving multiplex proteomic technologies and thus studies of single proteins were not included. Differences between proteomic technologies may introduce bias and warrant future investigation. For example, while most cancer proteins likely appear in the circulation from apoptosing cells, precancers are senescent and may produce protein below limits of detection across varying proteomic platforms. Further, meta-analyses were limited to available study-level rather than individual-level data and thus could not explore unmeasured or residual confounding within studies. Of note, the time between blood collection and diagnosis may influence observed associations, with results from studies with shorter time to diagnosis being more likely to reflect markers of existing cancer, and longer time to diagnosis more likely to reflect aetiological associations. Repeated protein measurements over time were not available within cohorts and represent opportunities to better investigate the change of precancer biomarkers over time. Disentangling sources of heterogeneity is challenging and highlights the need for a critical approach when bringing data together from different platforms. Such differences emphasize the value of large and mature prospective cohort studies with robust proteomic methods for precise, replicable, and clinically meaningful estimates.

This study represents the most comprehensive available evidence base to guide future research on circulating proteins associated with cancer risk. Plausible evidence was detected for protein-cancer-associations with several supported by complementary genetic evidence. The findings from this review show, first, the need for large and mature prospective proteomic cohort studies with high throughput methods and the importance of robust and replicable results. Second, the value of a critical approach when bringing together data from different platforms. Third, a lack of representation and diversity across global data and a paucity of data across several high-incidence cancer sites. Identifying proteins associated with cancer risk may better elucidate our understanding of cancer aetiology, as meaningful tools to inform cancer prevention and treatment.

## Data Availability

All data in the present systematic literature review and meta-analysis are available in the original published articles included in the review and upon reasonable request to the authors.

