## Supplementary data for "The circulating proteome and cancer risk: A systematic literature review and meta-analysis of 26 prospective studies with genetic validation"

### Supplementary Material

#### Supplementary Table 1. Search strategy for Medline

| Number | Search term |
|---|---|
| 1 | ((breast* or lung* or colon* or colorectal* or bowel* or rectum* or rectal or lower gi or prostat* or gastric* or stomach* or gastrointest* or liver* or hepat* or cervic* or cervix* or esophag* or oesophag* or upper gi or thyroid* or bladd* or urothel*) adj2 (cancer or carcinoma or adenocarcinoma* or neoplas* or malignan* or tumor* or tumour*)).mp. |
| 2 | (NSCLC* or SCLC* or UCC).mp. |
| 3 | or/1-2 |
| 4 | Incidence/ or inciden*.mp. |
| 5 | Risk/ or Risk Factors/ |
| 6 | risk*.mp. |
| 7 | detect*.mp. |
| 8 | predict*.mp. |
| 9 | diagnosis/ or Early detection of cancer/ or exp neoplasms/di |
| 10 | diagnos*.mp. |
| 11 | (prediagnos* or pre-diagnos*).mp. |
| 12 | or/4-11 |
| 13 | ((multiplex* or multi-plex or massspec* or mass-spec* or MS) and (protein* or proteom*)).mp. |
| 14 | (olink* or o-link*).mp. |
| 15 | (somalogic* or soma-logic* or somascan* or soma-scan*).mp. |
| 16 | luminex*.mp. |
| 17 | exp Neoplasm Proteins/bl |
| 18 | Proteome/ or Proteomics/ |
| 19 | or/13-18 |
| 20 | Epidemiologic studies/ |
| 21 | exp case control studies/ |
| 22 | exp cohort studies/ |
| 23 | case control.tw. |
| 24 | (cohort adj (study or studies)).tw. |
| 25 | Cohort analy$.tw. |
| 26 | (Follow up adj (study or studies)).tw. |
| 27 | (observational adj (study or studies)).tw. |
| 28 | Longitudinal.tw. |
| 29 | Prospective studies/ |
| 30 | exp Randomized Controlled Trial/ |
| 31 | 29 not 30 |
| 32 | biobank*.mp. |
| 33 | or/20-28,31-32 |
| 34 | 3 and 12 and 19 and 33 |

**Supplementary Table 2. Risk of bias assessment**

| Primary study | Selection | Comparability | Outcome/exposure | Total score |
|---|---|---|---|---|
| Albanes 2023 | ✸✸✸ | ✸✸ | ✸✸✸ | 8 |
| Aversa 2020 | ✸✸✸✸ | ✸✸ | ✸✸✸ | 9 |
| Bertuzzi 2015 | ✸✸✸✸ | ✸✸ | ✸✸✸ | 9 |
| Butt 2020 | ✸✸✸✸ | ✸✸ | ✸✸✸ | 9 |
| Camargo 2019 | ✸✸✸✸ | ✸✸ | ✸✸✸ | 9 |
| Chen 2017 | ✸✸✸ | ✸✸ | ✸✸✸ | 8 |
| Cook 2019 | ✸✸✸✸ | ✸✸ | ✸✸✸ | 9 |
| Epplein 2013 | ✸✸✸ | ✸ † | ✸✸✸ | 7 |
| Gaudet 2010 | ✸✸✸✸ | ✸ † | ✸✸✸ | 8 |
| Harlid 2021 | ✸✸✸✸ | ✸✸ | ✸✸✸ | 9 |
| Jakszyn 2017 | ✸✸✸✸ | ✸✸ | ✸✸✸ | 9 |
| Jovani 2022 | ✸✸✸✸ | ✸✸ | ✸✸✸ | 9 |
| Keeley 2014 | ✸✸✸✸ | ✸✸ | ✸✸✸ | 9 |
| Papier 2024 | ✸✸✸✸ | ✸✸ | ✸✸✸ | 9 |
| Shiels 2017 | ✸✸✸ | ✸ † | ✸✸✸ | 7 |
| Song 2018 | ✸✸✸✸ | ✸✸ | ✸✸✸ | 9 |
| Sun 2022 | ✸✸✸ | ✸ † | ✸✸✸ | 7 |
| Teras 2018 | ✸✸✸✸ | ✸✸ | ✸✸✸ | 9 |
| Wang 2023 | ✸✸✸✸ | ✸✸ | ✸✸✸ | 9 |
| Yoon 2022 | ✸✸✸✸ | ✸✸ | ✸✸✸ | 9 |
| Dagnino 2021 | ✸✸✸✸ | ✸✸ | ✸✸✸ | 9 |
| Wong 2011 | ✸✸✸ | ✸ † | ✸✸✸ | 7 |
| Ohishi 2013 | ✸✸✸ | ✸✸ | ✸✸✸ | 8 |
| Shiels, 2015 | ✸✸✸✸ | ✸✸ | ✸✸✸ | 9 |
| Ho, 2014 | ✸✸✸ | ✸ † | ✸✸✸ | 7 |
| Grassman, 2024 | ✸✸✸✸ | ✸ † | ✸✸✸ | 8 |

The maximum score for the Newcastle-Ottawa Scale for non-randomised studies is 9 stars, including 4 for selection, 2 for comparability, and 3 for outcome or exposure. Studies were graded as follows: Unsatisfactory = 0 to 3; Satisfactory = 4 to 5; Good = 6 to 7; Very good = 8 to 9. † Adjustment for sex was not applicable to single-sex studies.

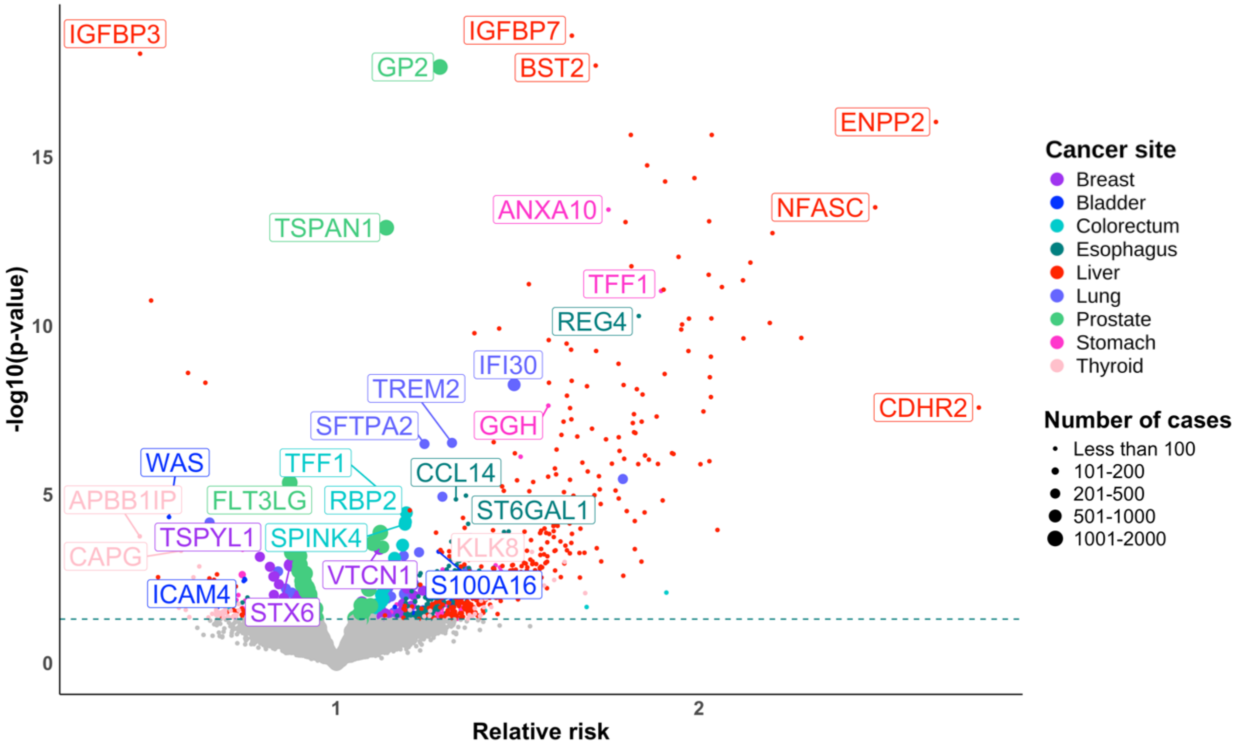


**Supplementary Figure 1. Protein-cancer-associations not included in meta-analyses due to reporting in single studies only.**

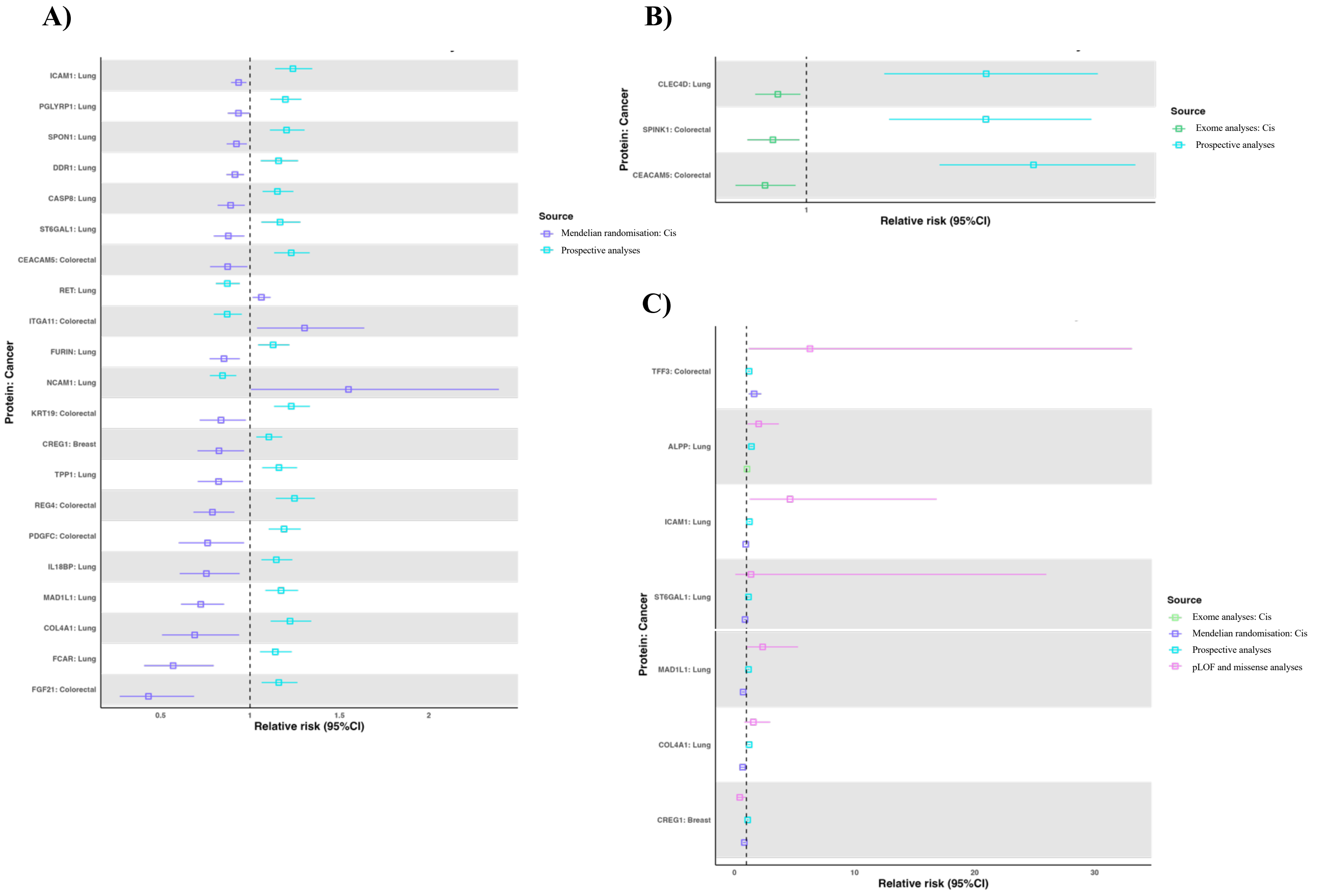


**Supplementary Figure 2. Genetic evidence for FDR-significant proteins identified in the meta-analysis of prospective data: Discordant findings compared to the observational analyses**

Mendelian randomisation (A) and exome (B) evidence; C) Probable loss-of-function (pLOF) and missense analyses evidence for protein-cancer-associations. **Abbreviations**: CI, confidence interval; FDR, false discovery rate; pLOF, probable loss-of-function. **Notes:** Relative risks (95% confidence intervals) for proteins identified in the meta-analyses of prospective data, for which genetic evidence was also available and directionally discordant associations were observed.

**Supplementary Methods**

We conducted two-sample Mendelian randomisation (MR) analyses to investigate potential causal associations of false discovery rate (FDR) significant circulating protein-cancer-associations identified in the meta-analyses of prospective data. Summary statistics for protein cis-protein quantitative trait loci (cis-pQTLs) were obtained from a genome-wide association study (GWAS) of 34,557 participants from the UK Biobank reported by Sun et al.[1] Genetic association estimates for cis-pQTLs and cancer endpoints were matched preferentially on rsID or by selecting a proxy single-nucleotide polymorphism (SNP) with the highest $r^2$ with the index cis-pQTL. Harmonised SNPs were oriented to the protein-increasing allele. MR estimates for each cis-pQTL and cancer pair were calculated using the Wald-ratio (β-cancer/β-protein) and expressed per SD increase in protein level. If multiple independent ($r^2 <0.01$) cis-pQTLs were associated with the same protein, the inverse-variance weighted (IVW) method was used to obtain combined associations with each cancer. Bonferroni correction was applied to account for multiple testing. MR associations that passed multiple-testing correction were then evaluated using colocalization analyses, to further determine whether the observed MR association was likely attributed to genetic confounding. We tested for the presence of a single shared causal variant between protein and cancer using conventional colocalization, as well as multiple shared causal variants using the Sum of Single Effects (SuSiE) regression method. Analyses were conducted in R (version 4.1.1) using TwoSampleMR and coloc R packages. We further investigated exome-wide genetic scores (exGS). Variants were oriented to the protein-increasing allele and exGS were calculated by summing the number of independent (clumping $r^2 <0.01$, 10,000KB) protein-increasing alleles, weighted by β reported by Dhindsa et al.[2] and projected in up to 337,543 UK Biobank participants with exome-sequencing using PLINK2.[3] We used logistic regression to estimate the association of each genetically predicted protein with cancer risk for each protein-cancer-association. Models were adjusted for age, sex, and the first ten genetic principal components of ancestry. Bonferroni correction was applied to account for multiple testing.

**PRISMA Checklist**

| Section and Topic | Item # | Checklist item | Location where item is reported |
|---|---|---|---|
| **TITLE** | | | |
| Title | 1 | Identify the report as a systematic review. | Page 4 |
| **ABSTRACT** | | | |
| Abstract | 2 | See the PRISMA 2020 for Abstracts checklist. | Page 2 |
| **INTRODUCTION** | | | |
| Rationale | 3 | Describe the rationale for the review in the context of existing knowledge. | Page 4 |
| Objectives | 4 | Provide an explicit statement of the objective(s) or question(s) the review addresses. | Page 4 |
| **METHODS** | | | |
| Eligibility criteria | 5 | Specify the inclusion and exclusion criteria for the review and how studies were grouped for the syntheses. | Page 4-5 |
| Information sources | 6 | Specify all databases, registers, websites, organisations, reference lists and other sources searched or consulted to identify studies. Specify the date when each source was last searched or consulted. | Page 4 |
| Search strategy | 7 | Present the full search strategies for all databases, registers and websites, including any filters and limits used. | Page 4-5, supplementary page 1 |
| Selection process | 8 | Specify the methods used to decide whether a study met the inclusion criteria of the review, including how many reviewers screened each record and each report retrieved, whether they worked independently, and if applicable, details of automation tools used in the process. | Page 4-5 |
| Data collection process | 9 | Specify the methods used to collect data from reports, including how many reviewers collected data from each report, whether they worked independently, any processes for obtaining or confirming data from study investigators, and if applicable, details of automation tools used in the process. | Page 4-5 |
| Data items | 10a | List and define all outcomes for which data were sought. Specify whether all results that were compatible with each outcome domain in each study were sought (e.g. for all measures, time points, analyses), and if not, the methods used to decide which results to collect. | Page 4-5 |
| | 10b | List and define all other variables for which data were sought (e.g. participant and intervention characteristics, funding sources). Describe any assumptions made about any missing or unclear information. | Page 4-5 |
| Study risk of bias assessment | 11 | Specify the methods used to assess risk of bias in the included studies, including details of the tool(s) used, how many reviewers assessed each study and whether they worked independently, and if applicable, details of automation tools used in the process. | Page 5, supplementary page 2 |
| Effect measures | 12 | Specify for each outcome the effect measure(s) (e.g. risk ratio, mean difference) used in the synthesis or presentation of results. | Page 5 |
| Synthesis | 13a | Describe the processes used to decide which studies were eligible for each synthesis (e.g. tabulating the study | Page 4-5 |

| Section and Topic | Item # | Checklist item | Location where item is reported |
|---|---|---|---|
| methods | | intervention characteristics and comparing against the planned groups for each synthesis (item #5)). | |
| | 13b | Describe any methods required to prepare the data for presentation or synthesis, such as handling of missing summary statistics, or data conversions. | Page 4-5 |
| | 13c | Describe any methods used to tabulate or visually display results of individual studies and syntheses. | Page 5 |
| | 13d | Describe any methods used to synthesize results and provide a rationale for the choice(s). If meta-analysis was performed, describe the model(s), method(s) to identify the presence and extent of statistical heterogeneity, and software package(s) used. | Page 5 |
| | 13e | Describe any methods used to explore possible causes of heterogeneity among study results (e.g. subgroup analysis, meta-regression). | Page 5 |
| | 13f | Describe any sensitivity analyses conducted to assess robustness of the synthesized results. | Page 5 |
| Reporting bias assessment | 14 | Describe any methods used to assess risk of bias due to missing results in a synthesis (arising from reporting biases). | Page 5 |
| Certainty assessment | 15 | Describe any methods used to assess certainty (or confidence) in the body of evidence for an outcome. | Page 5 |
| **RESULTS** | | | |
| Study selection | 16a | Describe the results of the search and selection process, from the number of records identified in the search to the number of studies included in the review, ideally using a flow diagram. | Page 6, figure 1 |
| | 16b | Cite studies that might appear to meet the inclusion criteria, but which were excluded, and explain why they were excluded. | Page 6 |
| Study characteristics | 17 | Cite each included study and present its characteristics. | Page 13-15, figure 2 |
| Risk of bias in studies | 18 | Present assessments of risk of bias for each included study. | Supplementary page 2 |
| Results of individual studies | 19 | For all outcomes, present, for each study: (a) summary statistics for each group (where appropriate) and (b) an effect estimate and its precision (e.g. confidence/credible interval), ideally using structured tables or plots. | Supplementary page 3-4, figure 3-5 |
| Results of syntheses | 20a | For each synthesis, briefly summarise the characteristics and risk of bias among contributing studies. | Page 6-9 |
| | 20b | Present results of all statistical syntheses conducted. If meta-analysis was done, present for each the summary estimate and its precision (e.g. confidence/credible interval) and measures of statistical heterogeneity. If comparing groups, describe the direction of the effect. | Page 6-9 |
| | 20c | Present results of all investigations of possible causes of heterogeneity among study results. | Page 6-9 |
| | 20d | Present results of all sensitivity analyses conducted to assess the robustness of the synthesized results. | Page 6-9 |
| Reporting | 21 | Present assessments of risk of bias due to missing results (arising from reporting biases) for each synthesis | Page 6-9, |

| Section and Topic | Item # | Checklist item | Location where item is reported |
|---|---|---|---|
| biases | | assessed. | supplementary page 2 |
| Certainty of evidence | 22 | Present assessments of certainty (or confidence) in the body of evidence for each outcome assessed. | Page 6-9, supplementary page 2 |
| **DISCUSSION** | | | |
| Discussion | 23a | Provide a general interpretation of the results in the context of other evidence. | Page 9-11 |
| | 23b | Discuss any limitations of the evidence included in the review. | Page 10-11 |
| | 23c | Discuss any limitations of the review processes used. | Page 10-11 |
| | 23d | Discuss implications of the results for practice, policy, and future research. | Page 10-11 |
| **OTHER INFORMATION** | | | |
| Registration and protocol | 24a | Provide registration information for the review, including register name and registration number, or state that the review was not registered. | Page 5 |
| | 24b | Indicate where the review protocol can be accessed, or state that a protocol was not prepared. | Page 5 |
| | 24c | Describe and explain any amendments to information provided at registration or in the protocol. | Page 5 |
| Support | 25 | Describe sources of financial or non-financial support for the review, and the role of the funders or sponsors in the review. | Page 5-6 |
| Competing interests | 26 | Declare any competing interests of review authors. | Page 5-6 |
| Availability of data, code and other materials | 27 | Report which of the following are publicly available and where they can be found: template data collection forms; data extracted from included studies; data used for all analyses; analytic code; any other materials used in the review. | Page 4-5, 12-15, figure 2-5, supplementary page 3-4 |

*From:* Page MJ, McKenzie JE, Bossuyt PM, Boutron I, Hoffmann TC, Mulrow CD, et al. The PRISMA 2020 statement: an updated guideline for reporting systematic reviews. BMJ 2021;372:n71. doi: 10.1136/bmj.n71. This work is licensed under CC BY 4.0. To view a copy of this license, visit https://creativecommons.org/licenses/by/4.0/
